# Efficacy of multiple micronutrient supplements during pregnancy on zinc status among pregnant women in Ghana: a randomized controlled trial

**DOI:** 10.64898/2025.12.18.25342593

**Authors:** K. Ryan Wessells, Seth Adu-Afarwuah, Charles D. Arnold, Harriet Okronipa, Anna Lartey, Kathryn G. Dewey, Christine M. McDonald

**Author notes:** **Corresponding author:** K. Ryan Wessells Department of Nutrition, University of California, Davis 1 Shields Ave. Davis, CA 95616.

## Abstract

**Background:** Multiple micronutrient deficiencies, including zinc deficiency, remain highly prevalent among pregnant women in many low- and middle-income countries and are associated with a multitude of adverse outcomes.

**Objective:** The objectives of this analysis were to assess the impacts of prenatal multiple micronutrient supplements (MMS) compared to iron and folic acid (IFA) supplements on maternal zinc status among pregnant women at 36 weeks gestation and assess associations with birth outcomes.

**Methods:** We analyzed plasma zinc concentrations at baseline (< 20 weeks gestation) and late pregnancy (36 weeks gestation) from a sub-set of pregnant women randomly assigned to receive either daily MMS (containing 30 mg/d zinc) or IFA supplements (n = 125/group) in the iLiNS DYAD trial in Ghana. Gestational and infant anthropometric outcomes were measured at birth.

**Results:** At baseline, mean ± SD plasma zinc concentrations were 61.9 ± 13.6 µg/dL and 16.5% of women were zinc deficient (plasma zinc concentration < 50 µg/dL). At 36 weeks gestation, mean ± SD plasma zinc concentrations were lower as compared to baseline, but did not differ significantly between groups (IFA: 52.2 ± 8.1 µg/dL vs. MMS: 52.9 ± 9.5 µg/dL; p = 0.25). Similarly, there was no difference in the estimated prevalence of zinc deficiency between groups at 36 weeks (IFA: 43.0% vs. MMS: 38.4%, p = 0.24). There were few significant associations between maternal plasma zinc concentrations or zinc deficiency and birth outcomes.

**Conclusions:** Daily MMS supplementation for ≥ 16 weeks during pregnancy did not significantly increase plasma zinc concentrations, and the prevalence of zinc deficiency increased over the course of pregnancy. Other complementary interventions to address both nutritional and underlying non-nutritional causes of zinc deficiency may be necessary to improve zinc status during pregnancy in this, and other vulnerable, populations.

**Registry:** The trial was registered as a clinical trial on ClinicalTrials.gov (NCT00970866) on September 2, 2009. https://clinicaltrials.gov/study/NCT00970866.

## Introduction

Multiple micronutrient deficiencies remain highly prevalent among pregnant women in many low- and middle-income countries (LMICs) and are associated with a multitude of adverse outcomes, including fetal growth restriction, preterm birth, low birth weight, and infant and maternal mortality (1–3). Although nationally-representative biochemical data on micronutrient status among pregnant women are limited, a recent analysis by Stevens et al. estimated that globally, 69% of non-pregnant women of reproductive age were deficient in at least one of three core micronutrients (e.g., iron, zinc, and folate) (4). Importantly, in 16 of 18 LMICs with available data, the proportion of women of reproductive age (WRA) with low plasma/serum zinc concentrations exceeded 20%, the threshold used to identify a public health problem (5).

Iron and folic acid (IFA) supplementation has been the existing standard of care for pregnant women in low- and middle-income countries (6). However, prenatal multiple micronutrient supplements (MMS) have been shown to improve maternal nutritional status and reduce the risk of adverse outcomes (e.g., preterm birth, stillbirth, low birthweight, and small-for-gestational age birth), as compared to IFA (1, 7). Thus, in 2020, the World Health Organization issued a new context-specific recommendation on MMS indicating that “antenatal multiple micronutrient supplements that include iron and folic acid are recommended in the context of rigorous research”(8). The WHO Guideline Development Group identified a need for more evidence on the efficacy of the dose of iron in MMS, compared with IFA (30 mg vs. 60 mg, respectively) (8), and research has recently been completed to address this knowledge gap (9). However, there is also remaining uncertainty regarding the optimal dose of zinc in MMS to improve maternal zinc status and prevent the risk of preterm birth to the greatest extent possible. In the Women First trial, investigators recently observed that 40-70% of pregnant women had low serum zinc concentrations (< 50 μg/dL) in the third trimester (10), despite receiving a daily supplement containing 15 mg of zinc.

Therefore, this analysis assessed the impacts of prenatal MMS (containing 30 mg zinc) compared to IFA supplements, on maternal zinc status (plasma zinc concentrations and the prevalence of zinc deficiency) among pregnant women at 36 gestational weeks (after ≥ 16 weeks of intervention). As a secondary objective, we assessed associations between maternal zinc status at baseline or 36 weeks gestation, and birth outcomes (e.g., preterm birth, small-for-gestational age, duration of gestation, etc.).

## Methods

### Study design, setting and participants

The current analyses are based on a sub-sample of pregnant participants enrolled in the iLiNS-DYAD Ghana trial (11, 12). The iLiNS-DYAD Ghana trial was designed as a randomized, partially double blind, individually randomized controlled trial that compared three micronutrient supplements, and targeted women during pregnancy (from ≤ 20 gestational weeks to delivery), and the first six months post-partum (12). The supplements provided during pregnancy were: (a) daily iron and folic acid (IFA); (b) daily multiple micronutrients (MMS), and (c) daily small-quantity lipid-based nutrient supplements (SQ-LNS) (12); the IFA and MMS study arms were fully blinded. Detailed descriptions of the overall study design and study protocol, as well as the effect of the interventions on multiple outcomes have been reported previously (11, 12). The present analyses address zinc-related perinatal outcomes of the trial in the IFA and MMS intervention arms only.

The trial was conducted from December 2009 – 2014 in the Yilo Krobo and the Lower Manya Krobo Districts in the Eastern Region of Ghana. Pregnant women were recruited from the antenatal clinics of 4 health care facilities in the study catchment area. Women were invited for screening if they met the following criteria: age ≥ 18 y, ≤ 20 weeks of gestation, planned to reside in the study catchment area for the duration of the study, and were willing to accept home visits and consume the study supplements. Women were excluded from the study if they presented with a history of a serious medical condition (i.e., if their antenatal cards indicated HIV infection, tuberculosis, asthma, epilepsy or any malignancy), had a known milk or peanut allergy, or were participating in another research study.

Eligible women were contacted at home to provide informed consent. Consent materials were presented, both written and orally, in the presence of an impartial witness. Informed consent was obtained from each participant prior to her recruitment in the study and documented as either a written signature or a fingerprint. Ethical approvals for the study were provided by the Ethical Review Committee of the Ghana Health Services, and the Institutional Review Boards of the University of Ghana Noguchi Memorial Institute for Medical Research and the University of California, Davis. The trial was registered as a clinical trial on ClinicalTrials.gov (NCT00970866).

### Group assignments and blinding

The study statistician at the University of California, Davis developed group allocations using a computer-generated block randomization scheme (SAS version 9.3; SAS institute) with block lengths of 9. Allocations were sequentially numbered 1 – 1320, placed in sealed, opaque envelopes and stacked in numerical order. After completing baseline assessments, recruited women who remained eligible for the study were enrolled; each enrolled participant picked one envelope from the 9 topmost envelops in the stack to reveal her group assignment (12). Group assignments for IFA and MMS were represented by 6 different color codes (3 for IFA, and 3 for MMS), and supplements were color-coded accordingly; investigators, study workers, participants and data analysts were blinded to the identities of the capsules. Allocation information was kept by the field supervisor and shared with the study statistician. Initial allocation codes were broken after the completion of all preliminary analyses. For the purpose of this analysis, intervention arms were randomly assigned novel blinded group codes, and all analyses were conducted and interpretation was agreed upon prior to unblinding group identity.

### Intervention products (intervention and comparator), trial supplements

During pregnancy, women randomized to the IFA group received a daily supplement containing 60 mg Fe and 400 µg folic acid; those randomized to the MMS group received a daily multiple micronutrient supplement containing 1-2 RDA of 18 vitamins and minerals, including 20 mg iron, 400 µg folic acid and 30 mg zinc (**Table 1**). The supplements were produced by DSM South Africa and provided as capsules packaged in blister packs. The formulation of the MMS supplements used in this trial were based on: (1) the UNIMMAP formulation (13), (2) a trial in Guinea Bissau, which showed a greater increase in birth weight among infants born to mothers supplemented with MMS containing twice the RDA of 15 micronutrients compared to those supplemented with MMS containing one RDA (14), and (3) a trial in Australia which indicated low dose iron supplements (20 mg) may be as efficacious in treating anemia in pregnancy as higher dose supplements, with fewer side effects (15).

**Table 1.** Micronutrient composition of supplements used in the study, with the United Nations International Multiple Micronutrient Antenatal Preparation (UNIMMAP) multiple micronutrient supplement product specification for comparison.

| Nutrient | IFA | MMS | UNIMMAP Formulation (13) |
| --- | --- | --- | --- |
| Vitamin A, µg RE |  | 800 | 800 |
| Vitamin B-1, mg |  | 2.8 | 1.4 |
| Vitamin B-2, mg |  | 2.8 | 1.4 |
| Niacin, mg |  | 36 | 18 |
| Pantothenic acid, mg |  | 7 |  |
| Vitamin B-6, mg |  | 3.8 | 1.9 |
| Folic acid, µg | 400 | 400 | 400 |
| Vitamin B-12, µg |  | 5.2 | 2.6 |
| Vitamin C, mg |  | 100 | 70 |
| Vitamin D, IU |  | 400 | 200 |
| Vitamin E, mg |  | 20 | 10 |
| Vitamin K, µg |  | 45 |  |
| Copper, mg |  | 4 | 2 |
| Iodine, µg |  | 250 | 150 |
| Iron, mg | 60 | 20 | 30 |
| Manganese, mg |  | 2.6 |  |
| Selenium, µg |  | 130 | 65 |
| Zinc, mg |  | 30 | 15 |

IFA, iron and folic acid; MMS, multiple micronutrient supplement; UNIMMAP, United Nations International Multiple Micronutrient Antenatal Preparation

### Procedures

At baseline, information on maternal and household socio-economic and demographic characteristics was collected via structured interview. Principal components analysis was used to derive an assets index, constructed based on ownership of a set of assets and household characteristics. Household food insecurity was assessed using the Household Food Insecurity Access Scale (HFIAS) (16). Gestational age was determined by ultrasound biometry (Aloka SSD 500, Tokyo, Japan), and anthropometric measurements were collected in duplicate for maternal height (Seca 217; Seca GmbH & Co, Hamburg, Germany), weight (Seca 874) and mid-upper arm circumference (Weigh & Measure, LLC, Olney, Maryland, USA). Venous hemoglobin concentration was determined using a Hemocue model 301 system (Hemocue AG, Wetzikon, Switzerland), and venous blood samples (7.5 ml) were collected into trace-element free, polyethylene tubes containing lithium heparin (Sarstedt AG & Co, Numbrecht, Germany).

At the end of the baseline visit, women received a 2-week supply of their assigned supplement and were instructed to consume one capsule daily with water after a meal. For the duration of the intervention (during pregnancy), all women were visited bi-weekly in their homes by a study fieldworker who was responsible for replenishing supplements, assessing adherence, and conducting a systematic symptom-based morbidity recall.

At 36 gestational weeks, women attended a secondary laboratory visit, where baseline anthropometric and biochemical assessments were repeated. Within 48 hours after birth, women were visited in their home or the hospital, and anthropometric measurements of their newborn were completed. Birth weight was measured to the nearest 20 g (Seca 383), and length (Seca 416), head and mid-upper arm circumferences (Weigh & Measure, LLC, Olney, Maryland, USA) to the nearest 0.1 cm. For infants, weight-for-age (WAZ), length-for-age (LAZ), BMI-for-age (BMIZ) and head circumference-for-age (HCAZ) z-scores were calculated according to the WHO growth standards (17, 18). Weight-for-gestational-age (WGAZ), length-for-gestational age (LGAZ) and head circumference-for-gestational-age (HCGAZ) z-scores were calculated according to Intergrowth 21^st^ standards (19). For newborns measured between 3-14 days (8.3% of sample), we back-calculated birth weight, length, and head circumference on the basis of z-scores at the time of measurement using the formulas described by the WHO, assuming z-scores remained constant (18).

### Sample processing and laboratory analyses

Venous blood samples were stored at 4°C until centrifugation, then centrifuged at 1252 x g for 15 minutes and plasma was aliquoted into microcentrifuge tubes. Plasma samples intended for nutritional biomarker assays were stored in Ghana at −20°C and later shipped on dry ice via air freight to the University of California, Davis where samples were stored at −70°C until analysis.

Plasma zinc was analyzed by inductively couple plasma optical emission spectrophotometry (ICP-OES 5900; Agilent Technologies, Inc., Santa Clara, CA, USA) at the University of California, San Francisco MLK Core Facility (Oakland, CA, USA). Details of the plasma zinc assay by ICP-OES have been described previously (20). In brief, plasma samples were centrifuged at 4000 x g for 15 minutes and digested overnight at 60°C in trace element grade 70% HNO_3_. This solution was then diluted to a final concentration of 5.5% HNO_3_ and centrifuged at 3000 x g for 10 minutes before analysis. Each batch of samples run on the ICP-OES was analyzed with Seronorm Trace Elements Serum L-2 (Sero AS, Billingstad, Akershus, Norway) as the reference material. All samples from a particular participant were analyzed in the same analytic run and 11% of samples were run in triplicate. The mean (± SD) inter-run coefficient of variation for triplicate samples was 5.5% ± 8.4%. C-reactive protein (CRP; mg/L) and α-1-acid glycoprotein (AGP; g/L) concentrations were determined by Cobas Integra 400 plus Automatic Analyzer (Roche Diagnostics Corp., Indianapolis, IN, USA).

### Outcome variables

In the present analysis, the outcome variables for the primary intervention effect objective are plasma zinc concentration, adjusted for inflammation (21), and the prevalence of zinc deficiency at 36 weeks gestation. The outcome variables for the secondary associational objective are duration of gestation (weeks), birth weight (g), and anthropometric z-scores at birth, based on WHO and Intergrowth 21^st^ growth standards (17–19).

### Sample size

In the present analysis, the main outcome variable was endline plasma zinc concentration. To detect treatment-related differences between intervention groups with an effect size of ≥ 0.36 SD, a sample size of 125 participants per study intervention group was necessary (α = 0.05, β = 0.20). Samples for the present analysis were randomly selected if they met the following criteria: (a) participant received the assigned supplement for the entire intervention period (12), and (b) baseline and 36 week biospecimen samples were collected, previously analyzed for acute phase protein concentrations (CRP and AGP) and had a plasma aliquot with sufficient volume archived at the University of California, Davis.

### Data analysis

A detailed statistical analysis plan was developed prior to analysis and is available online (22). All analyses were completed using a complete-case intention-to-treat approach (23). Descriptive statistics were calculated for all variables. Model assumptions were assessed (e.g. with Shapiro-Wilk tests for normality and assessments of linearity).

Plasma zinc concentrations were adjusted for elevated acute phase proteins, using a regression approach adapted from the BRINDA project (21). Specifically, if CRP and/or AGP were negatively associated with the outcome (p < 0.05) at a given time point, an adjustment by internal regression correction was applied using the inflammation biomarker(s) that showed a significant association with the outcome. Zinc deficiency was defined as plasma zinc concentrations (adjusted for inflammation) < 56 µg/dL in the first trimester and < 50 µg/dL in the second and third trimesters (21, 24).

To assess the effect of the intervention we followed the pre-post study design, analyzing continuous outcomes using ANCOVA models to estimate mean differences and binary outcomes using modified Poisson regression to estimate prevalence ratios controlling for baseline plasma zinc concentration in minimally adjusted models. To increase statistical power, we explored secondary analyses fitting adjusted models controlling for baseline covariates associated with the outcome at p < 0.10. Potential adjustment variables included baseline BMI, gestational age at enrolment, maternal age, maternal education, assets index, household food insecurity index, primiparity, season at enrolment, hemoglobin concentration at baseline, hemolysis of blood sample, time of day of blood collection, sample processing time and study duration. Exploratory effect modification was assessed for maternal age, primiparity, baseline plasma zinc status, gestational age at enrolment, and compliance with supplementation. This was conducted by including an interaction term in minimally adjusted models and significant interactions (p < 0.10) were further examined with post-hoc stratified analyses and/or regions of significance analyses.

The analysis of the secondary objective quantifying the associations between baseline and 36-week plasma zinc concentration and birth outcomes used multivariable linear regression for continuous outcomes and modified Poisson regression for binary outcomes. All models controlled for intervention arm and the baseline covariates listed above, except baseline plasma zinc concentration. Since the association between plasma zinc concentration and birth outcomes may have differed by intervention arm at the 36-week time point, we first tested an interaction term between intervention and zinc concentration. If the p-for-interaction was < 0.10 then we also tested associations separately by intervention arm.

All testing was two-sided and presented with 95% confidence intervals. Analyses were conducted in STATA v17 (StataCorp, LLC; College Station, Texas, USA).

## Results

### Participant characteristics

A total of 250 pregnant women, who provided baseline and 36-week plasma samples, are included in the present analyses. At baseline, women were 27.3 ± 5.3 years of age; 28% were primiparous and mean gestational age was 16.4 ± 3.0 weeks (**Table 2**). 40% were anemic (Hb < 110 g/L) and 42% presented with subclinical inflammation (CRP > 5 mg/L and/or AGP >1 g/L) (Table 2). Baseline, but not endline, plasma zinc concentrations were associated with CRP concentrations (rho = −0.17, p = 0.007) and baseline plasma zinc concentrations were adjusted accordingly; plasma zinc concentrations were not associated with AGP at either time point.

**Table 2.** Baseline characteristics of participants.

|  | IFA | MMS |
| --- | --- | --- |
| Participants, n | 125 | 125 |
| Age, years<sup>1</sup> | 27.3 ± 5.2 | 27.3 ± 5.4 |
| Formal education, years | 7.7 ± 3.5 | 7.2 ± 3.5 |
| Household asset index | 0.21 ± 0.96 | 0.21 ± 0.78 |
| Household moderately or severely food insecure, n (%) | 34 (27.2) | 29 (23.4) |
| Gestational age at enrollment, weeks | 16.4 ± 3.1 | 16.4 ± 2.8 |
| Primiparous, n (%) | 38 (30.4) | 33 (26.4) |
| BMI<sup>2</sup>, kg/m<sup>2</sup> | 24.5 ± 4.7 | 23.7 ± 5.8 |
| Underweight (<18.5 kg/m<sup>2</sup>), n (%) | 6 (4.9) | 9 (7.4) |
| Normal (18.5-25 kg/m<sup>2</sup>), n (%) | 71 (58.2) | 78 (63.9) |
| Overweight or obese (>25 kg/m<sup>2</sup>), n (%) | 45 (36.9) | 35 (28.7) |
| Hemoglobin, g/L | 112 ± 13 | 111 ± 11 |
| Anemia (< 110 g/L), n (%) | 50 (40.0) | 50 (40.0) |
| Elevated CRP (> 5 mg/L), n (%) | 55 (44.0) | 49 (39.2) |
| Elevated AGP (> 1 g/L), n (%) | 11 (8.8) | 5 (4.0) |

AGP, α-1-acid glycoprotein; CRP, C-reactive protein; IFA, iron and folic acid; MMS, multiple micronutrient supplement
<sup>1</sup>Values are means ± SD or n (%), unless otherwise specified.
<sup>2</sup>We estimated weight back to the 9th week of gestation using a restricted cubic spline model regressing baseline weight on gestational age at enrollment with 4 knots based on quintiles in the study's full dataset.

At baseline, the mean unadjusted plasma zinc concentration was 56.6 ± 12.5 µg/dL, and 36.6% had low plasma zinc concentrations (**Supplementary Table 1**). After adjusting for inflammation (CRP only), mean plasma zinc concentration was 61.9 ± 13.6 µg/dL and the prevalence of low plasma zinc concentrations was 16.5% (**Table 3**).

**Table 3.** Plasma zinc concentrations and the prevalence of zinc deficiency among pregnant women, at baseline and 36 gestational weeks, by intervention group^1^.

|  | IFA<br>(n = 121) | MMS<br>(n = 125) | Minimally adjusted<br>P-value |
| --- | --- | --- | --- |
| Plasma zinc, $\mu\text{g/dL}$<sup>2</sup> | | | |
| Baseline<sup>3</sup> | 63.8 ± 14.3 | 60.1 ± 12.6 | |
| 36 gestational weeks | 52.2 ± 8.1 | 52.9 ± 9.5 | 0.25 |
| Zinc deficiency, n (%)<sup>4</sup> |  |  |  |
| Baseline<sup>3</sup> | 17 (14.1%) | 24 (19.2%) |  |
| 36 gestational weeks | 52 (43.0%) | 48 (38.4%) | 0.24 |

IFA, iron and folic acid; MMS, multiple micronutrient supplement
<sup>1</sup>The IFA group received 60 mg iron and 400 $\mu\text{g}$ folic acid per day. The MMS group received 1-2 Recommended Dietary Allowances of 18 vitamins and minerals (including 30 mg zinc, 20 mg iron and 400 $\mu\text{g}$ folic acid per day).
<sup>2</sup> Values are means ± SD or n (%). Results are based on ANCOVA and Poisson regression models for continuous and categorical outcome variables, respectively. The minimally adjusted models control for baseline plasma zinc concentration. None of the additional potential covariates evaluated (baseline BMI, gestational age at enrolment, maternal age, maternal education, assets index, household food insecurity index, primiparity, season at enrolment, hemoglobin concentration at baseline, hemolysis of blood sample, time of day of blood collection, sample processing time and study duration) were associated with the outcomes at $p < 0.10$ , so results of the fully adjusted models do not differ from those of the minimally adjusted p-values.
<sup>3</sup>Baseline, but not endline, plasma zinc concentrations were associated with CRP concentrations ( $\rho = -0.17$ , $p = 0.007$ ); baseline plasma zinc concentrations were adjusted according to the BRINDA method for CRP only (21). Data not adjusted for baseline are presented in Supplementary Table 1.
<sup>4</sup>Plasma zinc deficiency is defined as a plasma zinc concentration $< 56 \mu\text{g/dL}$ in the first trimester ( $< 14$ weeks), and $< 50 \mu\text{g/dL}$ in the second or third trimesters ( $\geq 14$ weeks) (24).

### Effects of the intervention

Over the course of the intervention period in pregnancy (23.3 + 3.3 weeks), reported adherence to IFA and MMS were 75% and 77% of days, respectively, and did not differ by group (p = 0.77).

At 36-weeks gestation, there were no statistically significant differences in mean plasma zinc concentrations between women in the MMS intervention arm vs. the IFA arm (52.9 ± 9.5 µg/dL vs. 52.2 ± 8.1 µg/dL) in the minimally adjusted model (i.e., adjusted for baseline plasma zinc concentration) (p = 0.26; Table 3, Supplementary Table 1) or in the adjusted models (Table 3). Similarly, the prevalence of zinc deficiency did not differ between intervention arms at 36-weeks gestation (MMS vs. IFA, 38.4% vs. 42.6%, respectively, p = 0.24). Maternal age, parity, gestational age at enrollment, baseline plasma zinc deficiency and compliance with the assigned supplement did not modify the effect of the intervention on plasma zinc concentrations nor the prevalence of zinc deficiency at endline (**Supplementary Table 2**).

### Association between zinc deficiency and birth outcomes

In general, plasma zinc concentrations and zinc deficiency, at both baseline and 36-weeks gestation, were not associated with birth outcomes. Out of 64 comparisons, 2 were marginally significant (3.1% of comparisons; fewer than the 6.4% that would be expected by chance alone) (**Table 4**). Women who had zinc deficiency at baseline had, on average, infants with marginally higher BMIZ at birth than women who were not zinc deficient at baseline (−0.26 ± 1.0 vs. −0.55 ± 0.99, p = 0.067). In addition, higher plasma zinc concentrations at 36 weeks gestation were associated with a reduced risk of stunting at birth based on the Intergrowth-21^st^ standards (2.9% vs. 6.1%, p = 0.075).

**Table 4.** Associations between zinc deficiency, at baseline and 36 weeks, and birth outcomes (adjusting for treatment arm at 36 weeks); olling for all covariates^1^.

| Birth outcome | Low plasma zinc concentration | Normal plasma zinc concentration | P value<sup>2</sup> for continuous zinc | P value for binary zinc |
| --- | --- | --- | --- | --- |
| *Zinc status at baseline (adjusted for inflammation)<sup>3</sup>, n* | 41 | 208 |  |  |
| Birth weight (g) | 3111 (408) | 3043 (364) | 0.24 | 0.14 |
| Weight-for-age Z-score (WAZ)<sup>4</sup> | -0.42 (0.87) | -0.55 (0.82) | 0.22 | 0.19 |
| Weight-for-gestational age Z-score (WGAZ) | -0.44 (0.93) | -0.57 (0.88) | 0.14 | 0.35 |
| Low birth weight (< 2500 g) | 2.4% | 6.0% | 0.72 | 0.66 |
| Small for gestational age (< 10<sup>th</sup> percentile) | 17.1% | 20.6% | 0.92 | 0.99 |
| Length-for-gestational age Z-score (LAZ) | -0.62 (0.76) | -0.51 (0.94) | 0.61 | 0.87 |
| LAZ < -2 SD | 4.9% | 4.5% | 0.60 | 0.49 |
| Length-for-gestational age Z-score (LGAZ) | -0.62 (0.75) | -0.46 (0.98) | 0.45 | 0.46 |
| LGAZ < -2 SD | 2.5% | 4.5% | 0.54 | 0.63 |
| BMI-for-age Z-score (BMIZ) | -0.26 (1.0) | -0.55 (0.99) | 0.26 | 0.07 |
| Low BMIZ < -2 SD | 4.9% | 6.5% | 0.84 | 0.96 |
| Head circumference-for-age Z-score (HCAZ)<sup>5</sup> | -0.18 (1.02) | -0.34 (1.03) | 0.93 | 0.30 |
| Low HCAZ < -2 SD<sup>5</sup> | 4.9% | 4.5% | 0.66 | 0.78 |
| Head circumference-for-gestational age Z-score (HCGAZ) | 0.02 (0.94) | -0.16 (1.05) | 0.64 | 0.41 |
| Low HCGAZ < -2 SD | 0% | 3.5% | -- | -- |
| Duration of gestation, weeks | 39.7 (1.3) | 39.7 (1.4) | 0.49 | 0.50 |
| *Zinc status at endline (not inflammation adjusted), n* | 91 | 158 |  |  |
| Birth weight (g) | 3078 (348) | 3044 (387) | 0.29 | 0.34 |
| Weight-for-age Z-score (WAZ) | -0.49 (0.76) | -0.54 (0.88) | 0.33 | 0.48 |
| Weight-for-gestational age Z-score (WGAZ) | -0.52 (0.90) | -0.55 (0.89) | 0.72 | 0.59 |
| Low birth weight (< 2500 g) | 4.1% | 5.8% | 0.29 | 0.56 |
| Small-for-gestational age (< 10<sup>th</sup> percentile) | 22.5% | 18.7% | 0.44 | 0.35 |
| Length-for-gestational age Z-score (LAZ) | -0.56 (0.96) | -0.50 (0.89) | 0.93 | 0.62 |
| LAZ < -2 SD | 4.1% | 5.0% | 0.17 | 0.12 |
| Length-for-gestational age Z-score (LGAZ) | -0.52 (1.0) | -0.46 (0.9) | 0.63 | 0.66 |
| LGAZ < -2 SD | 6.1% | 2.9% | 0.08 | 0.11 |
| BMI-for-age Z-score (BMIZ) | -0.41 (0.98) | -0.55 (0.99) | 0.15 | 0.17 |
| Low BMIZ < -2 SD | 6.1% | 5.8% | 0.39 | 0.79 |
| Head circumference-for-age Z-score (HCAZ) | -0.29 (0.95) | -0.3 (1.1) | 0.84 | 0.84 |
| Low HCAZ < -2 SD | 6.1% | 2.9% | 0.18 | 0.31 |
| Head circumference-for-gestational age Z-score (HCGAZ) | -0.11 (0.99) | -0.12 (1.1) | 0.60 | 0.80 |
| Low HCGAZ < -2 SD | 3.1% | 2.9% | 0.99 | 0.72 |
| Duration of gestation, weeks | 39.8 (1.3) | 39.7 (1.4) | 0.32 | 0.70 |

BMIZ, BMI-for-age z-score; HCAZ, head circumference-for-age z-score; HCGAZ, head circumference-for-gestational age z-score; LAZ, length-for-age z-score; LGAZ, length-for-gestational age z-score; WAZ, weight-for-age z-score; WGAZ, weight-for-gestational age z-score
<sup>1</sup>Plasma zinc deficiency (low plasma zinc concentration) is defined as a plasma zinc concentration < 56 µg/dL in the first trimester (< 14 weeks), and <50 µg/dL in the second or third trimesters (≥ 14 weeks) (24). Normal plasma zinc concentration is defined as a plasma zinc concentration ≥ 56 µg/dL in the first trimester (< 14 weeks), and ≥ 50 µg/dL in the second or third trimesters (≥ 14 weeks).
<sup>2</sup>Values are means ± SD or n (%). Results are based on modified Poisson regression. All models include the following variables: baseline BMI, gestational age at enrolment, maternal age, maternal education, assets index, household food insecurity index, primiparity, season at enrolment, hemoglobin concentration at baseline, intervention arm, hemolysis of blood sample, time of day of blood collection, sample processing time and study duration.
<sup>3</sup>Baseline, but not endline, plasma zinc concentrations were associated with CRP concentrations (rho = -0.17, p = 0.007); baseline plasma zinc concentrations were adjusted according to the BRINDA method for CRP only (21).
<sup>4</sup>WAZ, LAZ, BMIZ, and HCAZ are calculated based on the WHO growth standards (17, 18). WGAZ, LGAZ, HCGAZ, and small for gestational age are calculated based on the INTERGROWTH 21<sup>st</sup> growth standards (19).
<sup>5</sup>P-for-interaction between continuous 36-wk plasma zinc concentration and intervention arm was assessed for all 16 outcomes. The indicated outcomes (2/16) had p-for-interaction <0.10 suggesting relationship between zinc and indicated outcome differed by intervention arm. Analyses stratified by intervention arm showed non-significant relationships (data not shown) and only full sample estimates are included here.

## Discussion

### Overview of the main results

In the present study, conducted among pregnant women in semi-urban communities in Ghana, the overall prevalence of low inflammation-adjusted plasma zinc concentrations increased over the course of pregnancy, from 16.5% to 40.5%, as measured at ∼16 and 36 gestational weeks, respectively. This suggests a high risk of zinc deficiency in the third trimester of pregnancy, and the potential of this population to benefit from zinc supplementation. However, there was no impact of the intervention on plasma zinc concentrations or the prevalence of zinc deficiency in this population. In addition, there were few significant associations between maternal plasma zinc concentrations or zinc deficiency and birth outcomes.

### Effects of the intervention

In the present study, plasma zinc concentrations did not respond to daily zinc supplementation (30 mg/d) provided as part of a multiple micronutrient supplement. The evidence on the effect of zinc supplementation (provided either alone, or as part of an MMS or SQ-LNS product) on zinc status in pregnancy is mixed. Our results are in contrast to a dose response meta-analysis of zinc supplementation in pregnancy, which reported that for the majority of studies included in the review, zinc supplementation (15-30 mg/d, provided as either zinc alone or MMS) significantly increased serum/plasma zinc status, or significantly reduced the number of low serum/plasma values in their participants (25). For every doubling of zinc intake, there was a significant 3% increase in serum/plasma zinc concentration among pregnant women (25). However, our results are in-line with a later meta-analysis, which reported that although zinc supplementation may have improved maternal serum/plasma zinc concentrations, the effect was not statistically significant (MD = 0.43 µmol/L; 95% CI −0.04 to 0.89; 5 studies), and that studies providing zinc only showed a greater increase (MD 0.86 µmol/L, 95% CI 0.67 to 1.05; n = 2) than studies that provided additional micronutrients (MD 0.01 µmol/L; 95% CI −0.72 to 0.72; n = 3) (26). In addition to the studies included in the aforementioned meta-analyses, several recent trials of zinc supplementation (providing 12-15 mg/d, in either MMS or SQ-LNS) have reported significant increases in plasma zinc concentrations (10, 27), whereas several other trials (providing 15 −30 mg Zn/d, as zinc + IFA or MMS supplements) have reported no effect of the intervention on zinc status (28–30). The observed lack of a consistent plasma zinc response to zinc supplementation during pregnancy, and consistently high prevalences of zinc deficiency remaining across all studies in spite of supplementation, may be due to (1) an inadequate amount of zinc in the MMS supplement, (2) inadequate absorption of zinc from the MMS supplement, and/or (3) non-nutritional causes of zinc deficiency.

In the present study, the MMS contained 30 mg Zn/d, which is almost 3 times the US FNB/IOM and IZiNCG RDA for zinc intake during pregnancy (11 mg/d and 13 mg/d, respectively) (24, 31) and twice the amount of zinc in the UNIMMAP formulation of MMS (13). Thus, it seems unlikely that the lack of response to zinc supplementation, and high prevalence of zinc deficiency even with supplementation, is due to an inadequate amount of zinc in the supplement. However, it is possible that pregnant women in the present study experienced poor zinc absorption from the supplement, in spite of expected enhanced zinc absorption in the third trimester of pregnancy to meet physiological needs (32). It is also possible that pregnant women may transfer more of the absorbed zinc to other tissues rather than increasing plasma zinc concentrations. Although we do not have individual level dietary data for participants, the study area’s predominantly maize-based diet is high in phytate, one of the main inhibitors of zinc absorption (33, 34). Second, the MMS supplement also included iron, which has been shown to impair zinc absorption (33, 35, 36). However, the ratio of iron: zinc was < 1:1 (20 mg iron, 30 mg zinc) in this supplement, which should limit the negative impact of iron on zinc absorption (37). Third, when supplemental zinc is consumed in the diet as a daily bolus dose, the fractional absorption of zinc has been shown to decrease (38). However, the amount of zinc provided by the MMS supplement (30 mg/d), in addition to dietary intake, should have been enough to meet physiological requirements for absorbed zinc in pregnancy (estimated range 2.27 – 5.02 mg/d) (24, 39, 40), even if the fractional absorption of zinc from the supplement was ≤15% (24). Finally, it is possible that there are non-nutritional causes of zinc deficiency in this population, leading to plasma zinc concentrations not being responsive to zinc supplementation in this population. At baseline, the estimated prevalence of zinc deficiency decreased by 20 percent points when controlling for inflammation (from 36% to 16%). Diarrheal diseases and intestinal infections increase zinc losses and other infections increase zinc requirements (e.g., malaria) (41, 42). At baseline, approximately 10% of pregnant women in the iLiNS-DYAD Ghana (parent) trial tested positive for malarial antigenemia (43), and 7% of women reported severe diarrhea and >33% of women reported fever and respiratory symptoms during each of the 2^nd^ and 3^rd^ trimesters of their pregnancies (43). Therefore, provision of zinc supplements may not be enough to control zinc deficiency; other strategies to address infection and inflammation may also be necessary.

### Associations between zinc status and birth outcomes

Two systematic reviews published in 2012 and 2015 demonstrated that preventive zinc supplementation resulted in a 14% reduction in preterm birth (RR: 0.86; 95% CI: 0.75, 0.99; and RR: 0.86; 95% CI: 0.76, 0.99, respectively) (12, 13) but had no detectable benefits on fetal growth. A more recent meta-analysis that included data from four new trials reported a similar 13% reduction in pre-term birth; however, this effect estimate did not reach statistical significance (RR: 0.87, 95% CI: 0.74, 1.03) (14); and again, there was no impact of the intervention on fetal growth. In addition, a recent meta-analysis by Smith et al. reported that MMS reduced the risk of preterm birth by 8% (RR: 0.92; 95% CI: 0.88-0.95) in comparison to IFA (8). However, in the iLiNS-DYAD Ghana (parent) trial, there was no impact of the intervention (MMS vs. IFA) on gestational age at birth or pre-term birth (12). However, there was a marginally significant reduction in the incidence of low birth weight among infants whose mothers received MMS as compared to IFA (12).

It is in this context that we examined associations between plasma zinc concentrations during pregnancy and birth outcomes. In the present study, however, there were no significant associations between plasma zinc concentrations, or zinc deficiency, at ∼16 and 36 weeks gestational and birth outcomes (e.g., birth weight, small-for-gestational age, stunting, etc.). Although we were unable to examine associations with pre-term birth, there were no significant associations between zinc status during pregnancy and duration of gestation. The literature on relationships between maternal plasma zinc concentrations and birth outcomes is mixed. In previous systematic reviews, approximately half of the studies reported significant associations between zinc status and birth outcomes, whereas the other half reported no significant associations (44, 45). For example, one study in Ethiopia (46) reported that 2^nd^ and 3^rd^ trimester plasma zinc concentrations were not significantly associated with low birth weight. However, another study in Ethiopia reported that plasma zinc concentrations were been positively correlated with birth weight (47). The Women First trial, conducted in the Democratic Republic of Congo, Pakistan, India and Guatemala observed that maternal plasma zinc concentrations were associated with birth weight and length, but not preterm birth (10).

### Strengths and weaknesses

This analysis had several strengths, including a fully randomized controlled design with an active control group, standardized biological sample collection and laboratory analyses based on procedures recommended by IZiNCG (48), and the analysis of samples for AGP and CRP to adjust for inflammation. However, this analysis also had several limitations. Adherence to supplementation was assessed by self-report and not direct observation, although participants were visited biweekly in their homes and the study team established a strong collaboration with the local antenatal health service providers. Secondly, plasma zinc concentrations are a relatively insensitive indicator of zinc nutritional status, as they are under homeostatic control. In addition, the cutoffs for plasma zinc concentrations during pregnancy used to indicate zinc deficiency account for hemodilution in pregnancy, they are based on limited data and do not account for time of day/fasting status (24). Therefore, although the high prevalence of low plasma zinc concentrations in this population appears to highlight a potential to benefit from zinc interventions, additional research is needed on cut-offs for plasma zinc concentrations indicative of zinc deficiency during pregnancy. Third, we do not have data on dietary zinc and phytate intake or biomarkers of environmental enteric dysfunction, which might help further elucidate underlying factors for the lack of responsiveness. Fourth, the sample size for these analyses was constrained by logistical and budgetary factors and was calculated to be able to detect an intervention effect of ≥ 0.36 SD; a larger sample size might have allowed us to detect a smaller intervention effect, although it may not have been clinically meaningful. Finally, because biological samples were collected at 36-weeks gestation, this sample does not include women who had a miscarriage or who gave birth prior to 36 weeks, which may introduce survivor bias and limit our ability to examine preterm birth as an outcome.

## Conclusions

In the present analysis, the estimated prevalence of zinc deficiency was high, but plasma zinc concentrations did not respond to a multiple-micronutrient supplement providing 30 mg Zn/d, which is twice the amount contained in UNIMMAP supplements. It appears that the 15 mg Zn contained in UNIMMAP supplements may not be adequate to improve plasma zinc concentrations in deficient populations. However, simply increasing the dose of zinc provided in MMS may also not be sufficient. Other complementary interventions to address underlying non-nutritional causes of zinc deficiency, including infection and inflammation control strategies, may be necessary to improve zinc status during pregnancy in vulnerable populations.

## Abbreviations

AGP: α-1-acid glycoprotein
BMIZ: body mass index z-score
BRINDA: Biomarkers Reflecting Inflammation and Nutritional Determinants of Anemia
CRP: C-reactive protein
FNB/IOM: Food and Nutrition Board Institute of Medicine
HCAGZ: head circumference-for-gestational age z-score
HCAZ: head circumference-for-age z-score
HFIAS: Household Food Insecurity Access Scale
ICP-OES: inductively coupled plasma ion exchange spectrometry
IFA: iron and folic acid
iZiNCG: International Zinc Nutrition Consultative Group
LAZ: length-for-age z-score
LGAZ: length-for-gestational age z-score
LMIC: low- and middle-income countries
MMS: multiple micronutrient supplement
SQ-LNS: small-quantity lipid-based nutrient supplements
UNIMMAP: United Nations International Multiple Micronutrient Antenatal Preparation
WAZ: weight-for-age z-score
WGAZ: weight-for-gestational age z-score
WRA: women of reproductive age

## Data Availability

Data described in the manuscript, code book, and analytic code will be made publicly and freely available without restriction at: https://osf.io/ek2gn

## Acknowledgements

We thank the laboratory staff Seth Antwi, Ronnie Osei Boateng, and Francis Maunger for their roles in the blood collection and plasma preparation; Marjorie Haskell at the University of California, Davis for biospecimen management; Lacey Baldiviez and Setti Shahab-Ferdows at the WHNRC for the CRP and AGP analyses; David Killilea and Kathleen Schultz at the University of California, San Francisco MLK Core Facility for the plasma zinc analyses; the iLiNS Project Steering Committee members Kenneth H. Brown, Mamane Zeilani, Stephen A. Vosti, and Jean Bosco Ouedraogo for advice in trial conceptualization; and Lindsay Allen for helping to define the SQ LNS formulation.

The authors’ responsibilities were as follows— SA-A, AL, and KGD: designed the research; SA-A, AL, and HO conducted the research; CDA: performed the statistical analysis; KRW and CMM: wrote the manuscript; SA-A, AL, HO, CDA and KGD: reviewed the draft manuscript; KRW: had primary responsibility for final content. All authors read and approved the final manuscript.

## Conflicts of Interest

All authors declare no conflicts of interest.

## Notes

**Sources of support:** Supported by Bill & Melinda Gates Foundation grant OPP49817 (to KGD) and a grant from the International Zinc Association (to CMM). Sponsors had no involvement or restrictions regarding publication.

### Competing Interest Statement

The authors have declared no competing interest.

### Clinical Trial

NCT00970866

### Funding Statement

This study was funded by a Bill & Melinda Gates Foundation grant OPP49817 (to KGD) and a grant from the International Zinc Association (to CMM).

### Author Declarations

The Ethical Review Committee of the Ghana Health Services, and the Institutional Review Boards of the University of Ghana Noguchi Memorial Institute for Medical Research and the University of California, Davis gave ethical approval for this work.

