## Supplementary Tables for "Efficacy of multiple micronutrient supplements during pregnancy on zinc status among pregnant women in Ghana: a randomized controlled trial"

Supplementary Table 1. Unadjusted plasma zinc concentrations and the prevalence of zinc deficiency among pregnant women, at baseline and 36 gestational weeks, by intervention group^1^

|  | IFA  (n = 121) | MMS  (n = 125) | Minimally adjusted P-value^2^ |
| --- | --- | --- | --- |
| Plasma zinc, µg/dL |  |  |  |
| Baseline^3^ | 58.1 ± 13.0 | 55.0 ± 11.9 |  |
| 36 gestational weeks | 52.1 ± 8.2 | 52.9 ± 9.5 | 0.28 |
| Zinc deficiency, n (%)^4^ |  |  |  |
| Baseline^3^ | 37 (30.6%) | 54 (43.2%) |  |
| 36 gestational weeks | 52 (43.0%) | 48 (38.4%) | 0.26 |

IFA, iron and folic acid; MMS, multiple micronutrient supplement

^1^The IFA group received 60 mg iron and 400 µg folic acid per day. The MMS group received 1-2 Recommended Dietary Allowances of 18 vitamins and minerals (including 30 mg zinc, 20 mg iron and 400 µg folic acid per day).

^2^ Values are means ± SD or n (%). Results are based on ANCOVA and Poisson regression models for continuous and categorical outcome variables, respectively. The minimally adjusted models control for baseline plasma zinc concentration. None of the additional potential covariates evaluated (baseline BMI, gestational age at enrolment, maternal age, maternal education, assets index, household food insecurity index, primiparity, season at enrolment, hemoglobin concentration at baseline, hemolysis of blood sample, time of day of blood collection, sample processing time and study duration) were associated with the outcomes at p<0.10, so results of the fully adjusted models do not differ from the minimally adjusted p-values.

^3^Plasma zinc concentrations are not adjusted for inflammation. Adjusted values are presented in Table 3.

^4^Plasma zinc deficiency is defined as a plasma zinc concentration < 56 µg/dL in the first trimester (< 14 weeks), and <50 µg/dL in the second or third trimesters (≥ 14 weeks) (24).

Supplementary Table 2. Effect modification testing of the impact of MMS vs. IFA on plasma zinc concentrations and the prevalence of zinc deficiency at endline

| Effect modifier | P-for-interaction value^1^ for continuous zinc | P-for-interaction value^1^ for binary zinc |
| --- | --- | --- |
| Maternal age (continuous) | 0.81 | 0.83 |
| Parity (primiparous vs multiparous) | 0.68 | 0.14 |
| Baseline zinc deficiency^2^ (zinc deficient vs normal) | 0.97 | 0.89 |
| Gestational age at enrollment (<14 vs. ≥ 14 weeks) | 0.54 | 0.23 |
| Compliance (<70% vs ≥ 70% supplement consumption) | 0.34 | 0.97 |

IFA, iron and folic acid supplement; MMS, multiple micronutrient supplement

^1^Results are based on ANCOVA and Poisson regression models for continuous and categorical outcome variables, respectively. Models control for baseline plasma zinc concentration and include an interaction term between effect modifier and intervention.

^3^Baseline, but not endline, plasma zinc concentrations were associated with CRP concentrations (rho = -0.17, p = 0.007); baseline plasma zinc concentrations were adjusted according to the BRINDA method for CRP only (21). Plasma zinc deficiency is defined as a plasma zinc concentration < 56 µg/dL in the first trimester (< 14 weeks), and <50 µg/dL in the second or third trimesters (≥ 14 weeks) (24).
